# Agentic Authoring of OMOP Concept Sets from Natural Language

**DOI:** 10.64898/2026.06.02.26354704

**Authors:** Hongyu Chen, Xing He, Hao Dai, Yu Huang, Mei Liu, Jiang Bian

## Abstract

Authoring OMOP concept sets from free-text descriptions remains a major bottleneck in scalable computable phenotyping for observational research. Existing tools support parts of this workflow but are designed primarily for interactive expert use rather than autonomous large language model (LLM) agents. We present an agentic framework that automatically generates OMOP concept sets by combining vocabulary tools, ontology extensions (RxClass, LOINC, and Disease Ontology), and procedural guidance. In ablation studies, the best configuration achieved Recall@100 of 0.965 and AP@100 of 0.875 on the development set. Cohort-level validation against OMOP-mapped EHR data yielded precision of 0.970, recall of 0.998, and a Jaccard index of 0.968. On an independent silver-standard benchmark of 457 concept-vocabulary pairs from 15 AD/ADRD target trial emulation studies, Recall@100 reached 0.835 and AP@100 reached 0.786. Task-specific tools outperformed unrestricted SQL access and PHOEBE 2.0, while progressive guidance performed best.

## Introduction

Real-world data captured in electronic health records (EHRs), administrative claims, and disease registries are increasingly used for observational research, comparative effectiveness studies, and regulatory decision-making. These studies require precise operational definitions of patient populations, exposures, outcomes, and covariates, typically implemented as clinical code sets — curated collections of standardized concepts that encode a clinical idea such as a disease, drug exposure, procedure, or laboratory measurement^1,2^. Regardless of the common data model (CDM), authoring these code sets remains a manual, iterative process that usually requires clarifying clinical intent, exploring synonyms and hierarchies, deciding which descendants to include, documenting exclusions, and validating the result against the local data source. In this work, we focus on concept sets within the Observational Health Data Sciences and Informatics (OHDSI) Observational Medical Outcomes Partnership (OMOP) CDM, where this challenge is particularly well defined because OMOP provides a unified vocabulary layer that makes concept sets both the standard authoring unit and a natural evaluation target.

Beyond time and expertise, manual concept-set authoring also undermines reproducibility. Prior work suggests that differences in codelists arise from population-specific design choices, database-specific validation history, expert judgment, temporal changes in coding practice, and broad-versus-narrow phenotype trade-offs^3^. Even when working within a CDM, independent teams attempting to implement the same observational study can produce materially different patient cohorts, in part due to the ambiguity of natural language study descriptions leave concept boundaries underspecified^4^. Repositories such as VSAC, OpenCodelists, PheKB, and the OHDSI Phenotype Library support reuse and transparency^2,5–7^, but reuse is rarely automatic. Concept sets often need to be adapted to the specific research question, design choices, data provenance, and vocabulary available in the analytic environment^8–10^.

A wide range of tools has been developed to reduce this burden. ATLAS, CodelistGenerator, and PHOEBE support candidate discovery through keyword search, hierarchy traversal, and recommendation workflows^11–14^. More recent systems extend automation in different directions: semi-automated codelist generation from trusted reference sources (GCAF^15^); embedding-based retrieval for ICD phenotypes (Phecoder^16^); LLM-driven cohort and feature extraction from database structure (EMR-AGENT^17^); automated ingestion and mapping of existing source code lists into ATLAS (CHIMERA^18^); MCP-based one-to-one medical concept standardization (Ahn et al.^19^); and LLM-based filtering of PHOEBE-recommended concepts (Anand et al.^20^). These systems improve important parts of the workflow, but each addresses a specific subtask — candidate discovery, code mapping, or concept filtering — rather than fully autonomous natural-language-to-concept-set authoring for observational studies.

End-to-end concept-set authoring is a multi-step reasoning process. Starting from a natural-language description, the author must decompose compound definitions into searchable components, assemble candidates from multiple vocabularies and domain-specific external resources, traverse hierarchies to capture descendants, resolve inclusions versus exclusions through explicit set subtraction, and judge whether the resulting set is semantically appropriate for the intended phenotype. Existing tools support individual steps in this pipeline but leave the sequencing, iteration, and stopping decisions to the user. LLM-based agentic systems with the ability to use tools offer a plausible architecture for this pipeline. Tools designed for agents can ground each operation—search, traversal, mapping, validation–using up-to-date vocabulary resources rather than relying on the LLM’s internal knowledge during training, while the agent handles sequencing, iteration, and stopping. However, tools alone specify only what the agent can do. Reliable authoring also requires explicit procedural knowledge—stepwise guidance that instructs the model how to choose, sequence, validate, and stop those operations.

In this work, we frame OMOP concept-set authoring as a knowledge-guided agentic task. We develop a framework that separates three design elements: an agentic controller that decides what to do and when, a structured tool/data layer that governs vocabulary and external-resource access, and a procedural knowledge layer operationalized as an agent skill. We evaluate the framework in two staged ablations: (1) one varying tool access from no vocabulary tools, to vanilla SQL access, to PHOEBE 2.0 wrapper, and finally to purpose-built structured tools with external resources extensions; and (2) under the best-performing tool configuration, one varying how the same procedural knowledge— guidance describing how the agent should use tools to accomplish the task—is delivered, comparing minimal guidance, static full guidance, and progressively on demand (i.e., providing relevant guidance only when the agent requests it). All configurations are assessed on a manually curated development set and an independent silver benchmark from Alzheimer’s disease and related dementias (AD/ADRD) target trial emulation studies using both concept-level retrieval metrics and downstream cohort-level impact.

## Methods

***Figure 1*** provides an overview of the study design. The system under evaluation is an LLM-based agentic framework that supports OMOP concept-set authoring from natural-language concept descriptions. Rather than treating the concept-set authoring as a single-step retrieval task, the framework is organized into three logical layers: (1) an a***gentic controller*** that reasons, calls tools, and iterates; (2) a ***structured tool/data layer*** that governs access to vocabulary and external resources; and (3) a ***procedural knowledge layer*** that specifies how the agent should carry out the authoring process. We evaluate these components in two staged ablations: a tool-access ablation and a knowledge-delivery ablation, while keeping the LLM backbone fixed unless otherwise noted. The following subsections describe the evaluation datasets, system architecture, tool/data layer, procedural knowledge layer, experimental conditions, evaluation metrics, and generalizability experiments.

**Figure 1.**
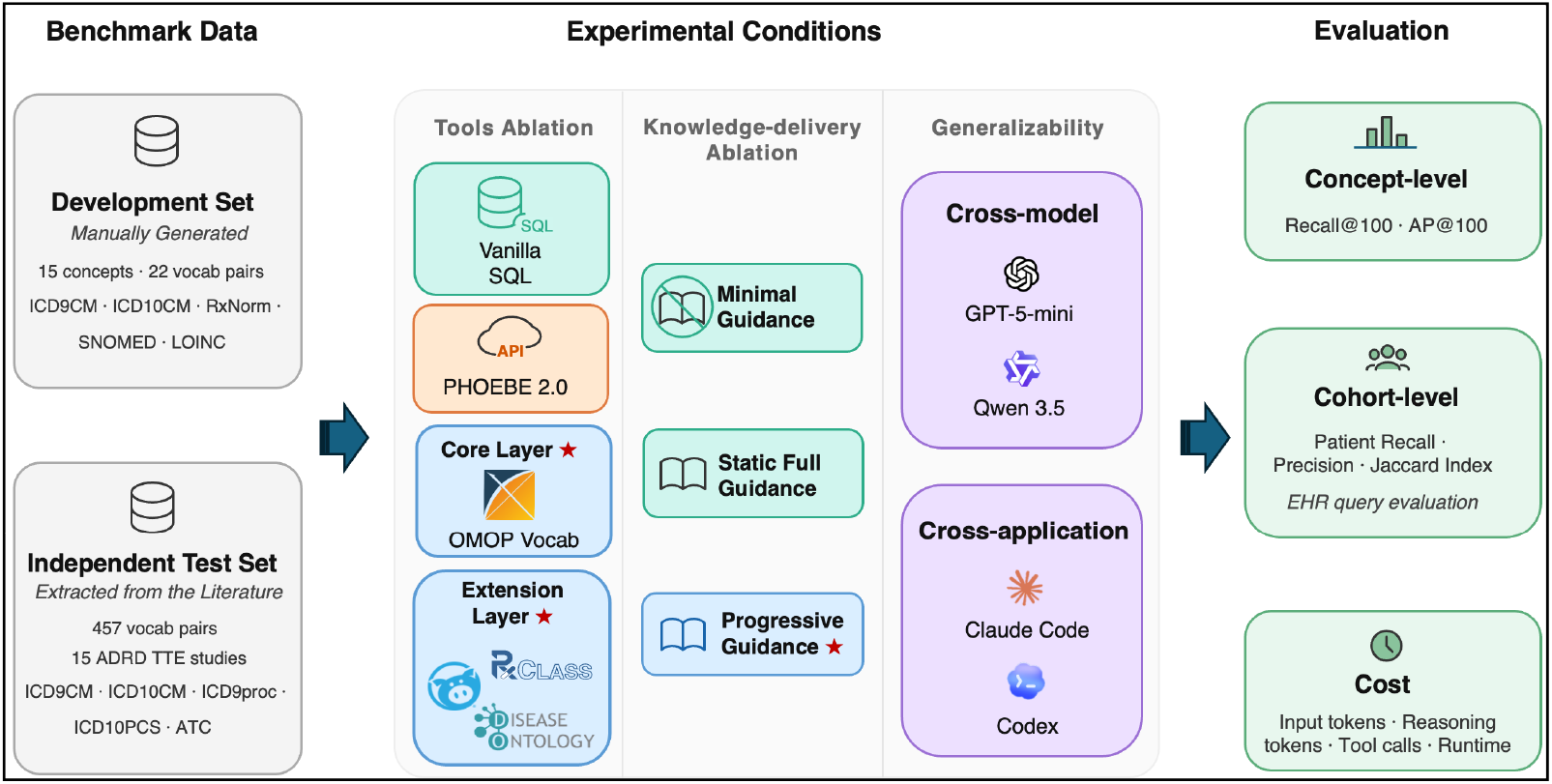
Study design. Stars indicate proposed modules.

### Data Sources and Annotation

Evaluating concept-set authoring requires high-quality reference concept sets and OMOP-formatted EHR datasets for downstream cohort-level validation. We used de-identified structured data from the University of Florida Health Integrated Data Repository covering January 2012 through May 2024. Data were restricted to patients with AD/ADRD or mild cognitive impairment, transformed to OMOP CDM v5.4, and used exclusively for cohort-level evaluation. The study was approved by the University of Florida Institutional Review Board (IRB #202001888).

#### Development set

We reused expert-curated concept sets from a prior AD/ADRD target trial emulation study conducted by our group. Each concept-vocabulary pair has been independently curated by two OMOP-experienced informatics experts and adjudicated to consensus. The development set contains 15 clinical concepts represented by 22 concept-vocabulary pairs across five vocabularies: ICD-9-CM (n = 7), ICD-10-CM (n = 7), RxNorm (n = 5), SNOMED CT (n = 2), and LOINC (n = 1).

#### Independent test set (silver benchmark)

To assess generalizability beyond a single study, we constructed a literature-derived benchmark from published AD/ADRD target trial emulation studies. We searched PubMed using the query (Alzheimer’s disease OR dementia OR cognitive impairment) AND (target trial emulation OR emulated trial) AND (real-world data OR electronic health records OR claims data), restricted to English-language original research published between 2018 and 2024 in first quartile (Q1) Journal Citation Reports (JCR)-ranked journals. We excluded case reports, reviews, editorials, and studies that did not report codelists using vocabularies supported by OMOP CDM.

Two reviewers independently extracted codelists from each retained study’s supplementary materials, appendices, or main text. For each reported code, reviewers (1) identified the source vocabulary (ICD-9-CM, ICD-10-CM, ICD-10-PCS, ICD-9-Proc, or ATC), (2) performed direct lookup in the OMOP vocabulary to obtain the corresponding concept_id, and (3) flagged codes that could not be unambiguously mapped (e.g., truncated codes [E11.*], version-ambiguous codes [ICD10], or codes not present in the OMOP vocabulary version used). Ambiguous mappings were resolved by consensus between reviewers with adjudication by a senior informatics expert when needed. Codes that remained unmappable after adjudication were excluded and documented. We term this a “silver” benchmark because, unlike the development set, these concept sets were not independently validated against a common EHR data source and reflect between-study variability in phenotype boundary decisions, vocabulary versions, and scope. The resulting benchmark contained 457 concept-vocabulary pairs from 15 studies across five vocabularies: ICD-10-CM (n=174), ICD-9-CM (n=87), ICD-10-PCS (n=3), ICD-9-Proc (n=3), and ATC (n=190).

### System Architecture

As illustrated in ***Figure 2***, the framework comprises three coordinated layers: an agentic controller, a structured tool/data layer, and a procedural knowledge layer. The agent receives a natural-language concept request together with a specification of the OMOP vocabularies from which candidate concepts should be drawn. It then reasons and executes iterative tool calls—interleaving vocabulary search, hierarchy traversal, cross-resource lookup, candidate validation, and gap analysis—and returns a ranked list of OMOP concept identifiers as the concept-set. The tool/data layer determines what retrieval and lookup operations are available; the procedural knowledge layer determines how the agent decomposes the request, selects retrieval routes, validates intermediate results, and decides when to stop. We implemented this procedural knowledge layer as a modular skill artifact, following the Anthropic Skills design framework^21^. The artifact specifies contextual information conditions for use (when the knowledge should be applied), task requirements, a generalized procedural outline, and domain-specific examples (playbooks) that support the agent’s decision-making during execution. Rather than exposing all guidance at once, the artifact reveals relevant instructions incrementally in response to the agent’s requests.

**Figure 2.**
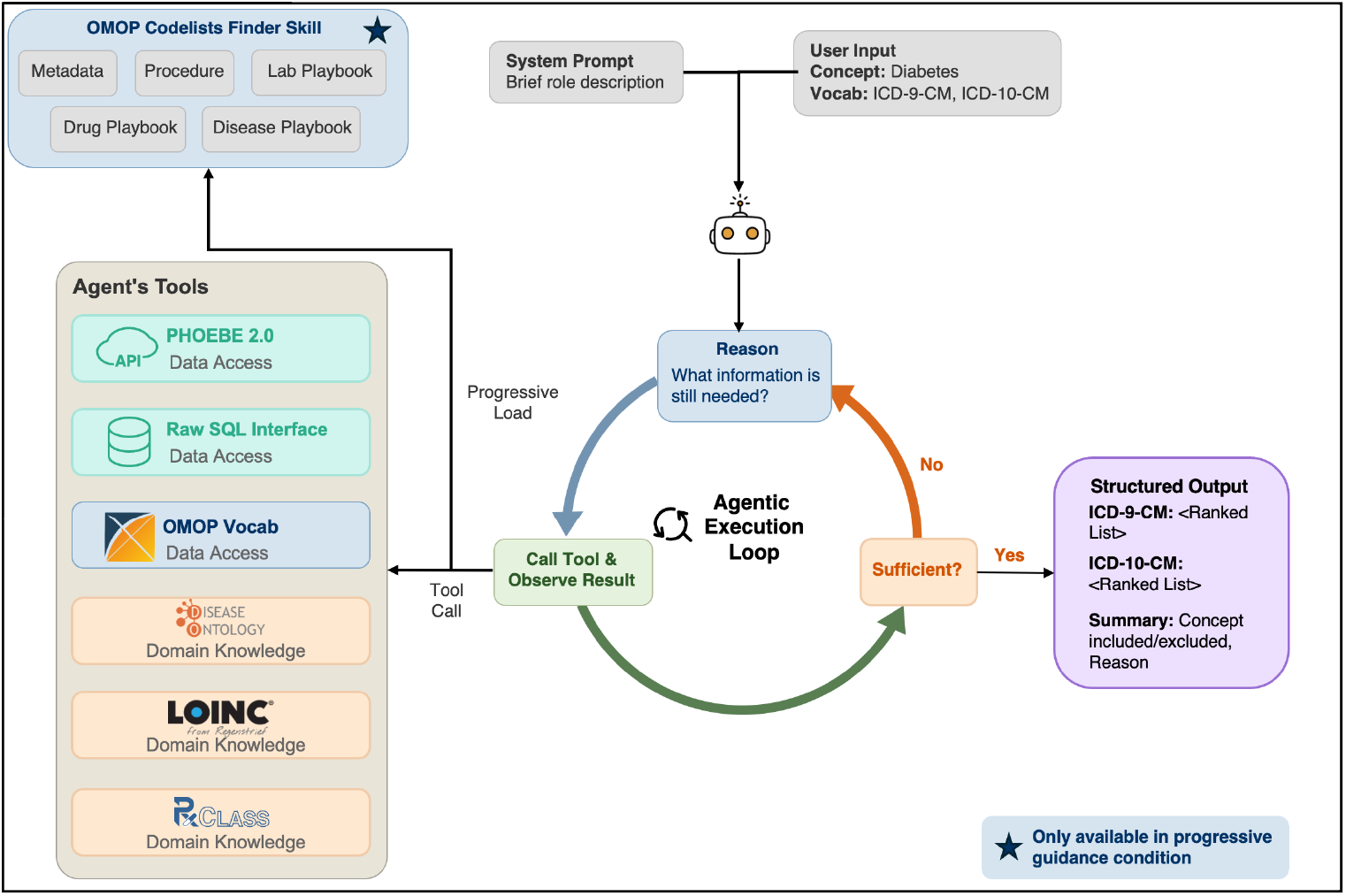
Overview of the agentic system.

### Tool/Data Layer

We designed the tool/data layer as a two-tier retrieval environment that exposes OMOP-native operations while selectively supplementing them with external resources.

#### Core layer

The core layer encapsulates the most common OMOP vocabulary operations into five parameterized tools, each corresponding to a single retrieval action: (1) concept name search, supporting wildcard matching with optional vocabulary and domain filters; (2) concept code search, supporting direct code lookup and prefix-based retrieval for hierarchical branches; (3) descendant expansion via the OMOP concept_ancestor table; (4) concept metadata retrieval, which returns domain, vocabulary, concept code and concept name; and (5) cross-vocabulary mapping through OMOP concept relationships. All five tools use predefined SQL templates, so the agent specifies retrieval intent through parameters rather than constructing free-form queries.

#### Extension layer

The extension layer adds targeted external resources to enrich retrieval where OMOP native structure is limited. For drugs, an NLM RxClass toolkit resolves class-to-ingredient membership through class search, member retrieval, and hierarchy traversal. For measurements, a LOINC toolkit provides panel membership and synonym relationships richer than the subset represented in OMOP. For conditions, a Disease Ontology toolkit supports concept expansion and cross-ontology mappings beyond OMOP-native links. These resources remain part of the tool/data layer because they expand what the agent can retrieve, not how the agent should reason about the task.

### Procedural Knowledge Layer

Tools expose capabilities but do not specify how concept-set authoring should proceed. To address this, we implemented a procedural knowledge layer as an agent skill (i.e., OMOP Codelists Finder Skill) that teaches the agent how to convert a free-text concept request into a high-coverage OMOP concept set. This layer comprises four components: (1) metadata describing when the skill should be loaded; (2) a core task contract specifying the expected outputs, validation targets, and stopping rules; (3) a workflow scaffold covering concept decomposition, synonym expansion, domain-aware retrieval, candidate validation, gap analysis, and final assembly; and (4) domain-specific playbooks for drugs, conditions, and measurements that encode vocabulary-specific retrieval pathways, exclusion logic, and common failure modes. In this Progressive Guidance condition, these components are stored separately; the metadata are injected into the system prompt, while the rest are loaded only when requested by the agent. This design allows simple concepts to be handled with the task contract and tools alone, whereas complex concepts trigger loading of the workflow scaffold and the relevant domain-specific playbook.

### Experimental Conditions

We used a two-stage ablation rather than a full factorial design. Stage 1 varied tool access while holding the agent architecture and model backbone fixed. Stage 2 fixed the best-performing tool configuration and varied how the same procedural knowledge was delivered.

#### Tool-access conditions

We evaluated five tool-access conditions organized into two groups. **Group A** contained three baseline conditions without custom tooling. **No Tool** provided the agent with no vocabulary access, testing whether LLM pretraining knowledge alone was sufficient. **Vanilla SQL** granted unrestricted SQL access to the full OMOP vocabulary database with no pre-built wrappers, requiring the agent to formulate every query itself. **PHOEBE 2.0** connected the agent to the OHDSI ATLAS WebAPI, enabling vocabulary search, recommender-based expansion, and cross-vocabulary mapping. Because PHOEBE is designed for interactive human review rather than autonomous multi-step execution, it returns broad recommendations. To adapt it for agentic use with only minimal adjustments, we paginated results and maintained intermediate outputs in seed and candidate pools to adjust for context limits. The seed pool maintained a running list of relevant concepts from any vocabulary, which were supplied to the agent in subsequent recommendation steps, while the candidate pool contained only concepts belonging to the vocabularies specified by the user. Aside from enforcing page-wise evaluation and instructing the agent to iteratively refine these pools, we did not implement a PHOEBE-specific agent strategy optimized for its recommender interface (e.g., tailored prompt engineering or context compression). Our results therefore represent PHOEBE operating under a generic agentic operating mode rather than its theoretical optimum. Developing and evaluating such a strategy is a direction for future work. **Group B** contained two purpose-built configurations. **OMOP Vocabulary Tools** exposed the five core-layer operations and established the structured-access baseline. **OMOP Vocabulary Tools + Extension Layer** added the RxClass, LOINC, and Disease Ontology toolkits to support enhanced retrieval for drugs, measurements, and conditions.

#### Knowledge-delivery conditions

Under the best-performing tool configuration, we compared three modes of delivering the same procedural knowledge. **Minimal Guidance** provided only the task contract and stopping rule, without an explicit workflow scaffold or domain-specific playbooks. **Static Full Guidance** loaded the complete procedural knowledge layer for every concept, including the full workflow scaffold and all domain-specific playbooks regardless of concept complexity. **Progressive Guidance** stored these components separately and loaded only those relevant to the current concept. This condition operationalizes progressive disclosure by keeping concise guidance always available while introducing detailed procedures and domain knowledge only when requested. This design allowed us to evaluate not only whether procedural knowledge helps, but also whether its selective delivery matters.

### Evaluation

We evaluate performance from three complementary perspectives.

▪ **Concept-level evaluation**. The primary metric was Recall@100, defined as the proportion of reference concepts recovered within the top 100 ranked candidates. Average Precision@100 (AP@100) served as the secondary metric, capturing both retrieval completeness and ranking quality. We used k = 100 to align with recent retrieval-style concept-authoring evaluations^16^. Metrics were computed per concept-vocabulary pair and macro-averaged across pairs. Concept-level evaluation was performed on both the development set (22 pairs) and the independent test set (457 pairs).
▪ **Cohort-level evaluation**. For each generated concept set, we queried the UF Health OMOP database to identify each patient’s first recorded occurrence of any concept in the set and compared the resulting patient cohort with the cohort generated from the expert-curated reference concept set. We report macro-averaged cohort precision, recall, and Jaccard index across the 22 development-set pairs. Cohort-level analysis was limited to the development set because patient-level EHR data were not available for the silver benchmark.
▪ **Cost evaluation**. To characterize computational overhead, we recorded average number of input tokens, reasoning tokens, tool calls, and wall-clock runtime per concept-vocabulary pair on the development set.

### Generalizability Experiments

We assessed whether the proposed tools and skill generalized across LLM backbones and end-user agentic applications using the best-performing configuration: OMOP Vocabulary Tools + Extension Layer with progressive skill loading.

▪ **Cross-model evaluation**. We repeated the full development-set evaluation with Qwen 3.5 397B A17B without modifying the tools or skill.
▪ **Cross-application evaluation**. To assess deployment feasibility, we exposed the toolkit as an MCP server and evaluated the same skill in Claude Code and the OpenAI Codex desktop application. We selected one representative concept per target vocabulary and recorded the retrieval pathway, resulting concept set, and Recall@100 against the development-set reference for each case.

### Experimental Settings

The primary agent was implemented in Python 3.12 with NumPy, pandas, and LangChain for orchestration. It was initialized using LangChain’s create_agent function, which creates agents following the ReAct (reasoning + acting) pattern^22^. A system prompt defining its role as an OMOP concept author was provided at initialization. The OMOP vocabulary was hosted in a local PostgreSQL 18.1 database, and inference was served through OpenRouter. Unless otherwise noted, all experiments used GPT-5-mini^23^ with high reasoning effort. Cross-model evaluation used Qwen 3.5 397B A17B^24^. Cross-application evaluation used Claude Sonnet 4.6 with Claude Code deployment (v1.1.4173) and GPT-5.3-Codex OpenAI Codex desktop application (v26.217.1959).

## Results

### Tool-Access Ablation

***Table 1*** summarizes concept-level performance across the five tool configurations on the development set. Across tool-access conditions, the largest performance gains came from moving from unstructured or weakly structured access to purpose-built retrieval tools. Although Vanilla SQL exposed the same underlying OMOP vocabulary data, it required the agent to independently formulate every query and traversal step, resulting in avoidable failures. By contrast, the structured OMOP tools parameterized the most common authoring operations around the vocabulary’s native structure, allowing the agent to focus on retrieval intent rather than query construction. The extension layer produced further gains, especially where OMOP alone does not encode the relationships needed for high-quality authoring, for example, class-to-ingredient reasoning.

**Table 1.** Concept-level tool ablation performance on the development set.

| Condition | PHOEBE 2.0 | Vanilla SQL | OMOP Vocab Tools | RxClass | LOINC | Disease Ontology | Recall @100 | AP@100 |
| --- | --- | --- | --- | --- | --- | --- | --- | --- |
| No Tool |  |  |  |  |  |  | 0.023 | 0.023 |
| Vanilla SQL |  | ✓ |  |  |  |  | 0.694 | 0.394 |
| PHOEBE 2.0 | ✓ |  |  |  |  |  | 0.179 | 0.084 |
| OMOP Vocab Tools |  |  | ✓ |  |  |  | 0.934 | 0.705 |
| OMOP + Extension Layer |  |  | ✓ | ✓ | ✓ | ✓ | **0.939** | **0.822** |

**Table 2.** Concept-level performance and cost for knowledge-delivery ablation conditions on the development set. All conditions use OMOP Vocabulary Tools + Extension Layer.

| Condition | Recall@100 | AP@100 | Input Tokens | Reasoning Tokens | Tool Calls | Runtime (s) |
| --- | --- | --- | --- | --- | --- | --- |
| Minimal Guidance | 0.939 | 0.822 | 29,667 | 5,155 | 159 | 185 |
| Static Full Guidance | 0.954 | 0.851 | 57,031 | 6,243 | 162 | 353 |
| **Progressive Guidance** | **0.965** | **0.875** | 34,685 | 5,461 | 158 | 221 |

The No Tool condition achieved Recall@100 of 0.023, confirming that internal knowledge alone was insufficient for reliable OMOP concept retrieval. Vanilla SQL improved Recall@100 to 0.694, but this remained far below the structured-tool configurations even though both conditions used the same underlying OMOP vocabulary database. The gap suggests that unstructured database access forces the agent to spend substantial reasoning effort on schema navigation and query construction rather than on the clinical retrieval problem itself.

PHOEBE 2.0 achieved Recall@100 of 0.179 and AP@100 of 0.084, substantially lower than Vanilla SQL. The predominant failure mode was drug class coverage: PHOEBE returned no candidate concepts for five drug-class pairs in the development set. Reasoning trace review suggested diminishing returns across rounds. After the first recommendation round, the agent rarely uncovered additional relevant concepts but continued to accumulate marginal candidates, which consumed context-window budget and complicated downstream selection. We note that this result reflects PHOEBE under a generic agentic wrapper rather than an optimized agent strategy tailored to its recommender interface. An optimized strategy—for example, more granular instructions to limit marginal concepts, mechanisms to merge similar concepts in the seed, or a multi-agent architecture to reduce per-agent context load —might yield materially different performance. The comparison should therefore be interpreted as a test of autonomous executability under standardized conditions, not a head-to-head benchmark of PHOEBE’s inherent recommendation quality.

OMOP Vocabulary Tools achieved Recall@100 of 0.934 and AP@100 of 0.705. Adding the extension layer increased Recall@100 modestly to 0.939 but improved AP@100 more substantially, from 0.705 to 0.822, indicating better ranking of relevant concepts. The benefit was concentrated in drug-class tasks: for RxNorm pairs, RxClass increased Recall@100 from 0.940 to 0.972 and AP@100 from 0.893 to 0.948. In contrast, LOINC and Disease Ontology contributed only incremental improvements in this AD/ADRD-focused evaluation set.

### Knowledge-Delivery Ablation

Structured retrieval tools alone established a strong baseline (Recall@100 = 0.939), indicating that much of concept-set authoring can be handled through well-designed retrieval actions combined with the model’s internal clinical knowledge. Additional gains were achieved through procedural knowledge, but the magnitude of these gains depended on how the knowledge was delivered. Static Full Guidance improved recall but incurred the largest context and runtime cost. Progressive Guidance achieved the best overall performance (Recall@100 = 0.965, AP@100 = 0.875) while using substantially fewer input tokens and less runtime than Static Full Guidance. This pattern suggests that selectively loaded knowledge is more effective than universally loaded instructions.

The added value of procedural knowledge was concentrated in concepts that required exclusions, multi-hop retrieval, or cross-resource reasoning. Examples included compound condition definitions such as AD/ADRD, which require hierarchical assembly, and “glucose-lowering drugs excluding insulin, GLP-1 receptor agonists, SGLT2 inhibitors”, containing logical operators. For such concepts, procedural knowledge primarily helps by routing the agent toward the appropriate retrieval pathway, prompting explicit gap analysis, and forcing validation before termination, rather than by compensating for weak tool access.

### Cohort-level Performance

***Table 3*** summarizes cohort-level performance, which broadly preserved the same ordering observed at the concept level, indicating that improved concept retrieval translated into more accurate patient cohort construction. The best configuration—Progressive Guidance with OMOP + Extension Layer—achieved cohort precision of 0.970, recall of 0.998, and Jaccard of 0.968. PHOEBE achieved cohort precision of 0.411, recall of 0.312, and Jaccard of 0.232. Vanilla SQL achieved cohort precision of 0.942 and recall of 0.906 despite its lower concept-level Recall@100 of 0.694, suggesting that it often recovered high-prevalence codes even when it missed less common supplementary concepts.

**Table 3.** Cohort-level performance on the development set.

| Condition | Precision | Recall | Jaccard |
| --- | --- | --- | --- |
| PHOEBE | 0.411 | 0.312 | 0.232 |
| Vanilla SQL | 0.942 | 0.906 | 0.893 |
| Minimal Guidance | 0.944 | 0.968 | 0.912 |
| Static Full Guidance | 0.846 | 0.994 | 0.843 |
| **Progressive Guidance** | **0.970** | **0.998** | **0.968** |

Static Full Guidance produced lower cohort precision (0.846) than the Minimal Guidance condition (0.944) despite higher concept-level Recall@100, indicating a cohort-level precision-recall tradeoff. Progressive Guidance provided the best overall balance across both concept-level and cohort-level metrics, reinforcing the advantage of selective knowledge delivery.

### Error Analysis

Reasoning-trace review identified four recurring error modes: model laziness, ambiguous concepts, misuse of knowledge, and random omissions. Examples of each error mode are provided in the GitHub repository (https://github.com/hongyuchen1/OMOP_Codelists_Finder). ***Figure 3*** shows the distribution of error modes across guidance conditions.

▪ **Model Laziness**. The agent occasionally terminated early or underused tools, relying excessively on internal knowledge rather than consulting external resources. Because internal knowledge may be incomplete or outdated, this behavior can result in missing relevant concepts. For example, in the AD/ADRD, missed descendants included Pick’s disease, age-related cognitive decline, and senile degeneration of the brain, which were not captured by the models’ internal knowledge, but Disease Ontology can capture these descendants.
▪ **Ambiguous Concepts**. Prior studies have noted that the conceptual boundary of a clinical idea is difficult to define even among domain experts^3,25^. In the AD/ADRD case, the agent included codes such as dementia with behavioral disturbance, or substance-induced dementia. Whether these should be included in an AD/ADRD concept set varies across studies. When the input description did not clearly define the intended boundary, the agent tended to adopt an inclusive interpretation. This discrepancy suggests that additional research is needed to explore implicit constraints beyond the concept itself.
▪ **Misuse of Knowledge**. The agent was encouraged to identify coding patterns in order to expand candidate concepts. However, this strategy occasionally led to incorrect generalization. For example, the agent correctly inferred from ICD-10-CM that codes under E11.* correspond specifically to Type 2 diabetes but incorrectly applied a similar pattern to ICD-9-CM, where 250.* includes both Type 1 and Type 2 diabetes. This mistake preserved recall but significantly reduced ranking precision.
▪ **Random Omissions**. Across conditions, the agent sometimes retrieved correct concepts earlier in the workflow but failed to include them in the final output. These omissions did not appear to be deliberate removals. This pattern was most visible in concepts with many descendants and is consistent with increasing context length making it more difficult for the agent to maintain a stable output.

**Figure 3.**
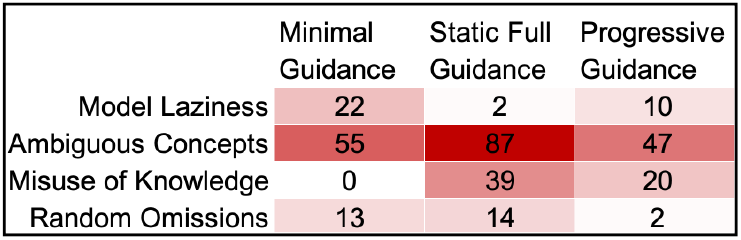
Distribution of error modes across guidance conditions. The cells are filled with the relative frequency of each error mode in each condition.

Taken together, these patterns help explain the ablation results. Under Minimal Guidance, the dominant failure modes were model laziness and ambiguity-related inclusion errors. Under Static Full Guidance, model laziness was largely suppressed, but this improvement came with a marked increase in ambiguity-related errors, misuse of knowledge, and random omissions. This shift suggests that supplying all procedural knowledge simultaneously reduced tool underuse but increased overgeneralization and context-related instability. In addition, the agent tends to incorporate borderline concepts, which further increase the errors related to ambiguous concepts. Progressive Guidance produced the most balanced error profile: although some model laziness remained, it had the fewest ambiguity-related errors and the fewest random omissions, while also reducing knowledge misuse relative to Static Full Guidance. Taken together, these patterns suggest that progressive disclosure improved performance not simply by adding more guidance, but by changing when and how procedural knowledge was introduced during execution. However, ambiguity in the definition of AD/ADRD remained a persistent challenge across all conditions and remains a major source of error and requires further study to collect study-specific information to disambiguate the concept.

### Generalizability

***Table 4*** demonstrates the generalizability of the best configuration across models, applications, and a literature-derived benchmark without model- or platform-specific redesign.

▪ **Cross-model**. Qwen 3.5 397B A17B achieved Recall@100 of 0.921 and AP@100 of 0.851 on the development set, retaining 95.4% of the GPT reference Recall@100 (0.965). This modest drop suggests that the tool interface and skill transfer reasonably well across LLM backends.
▪ **Cross-application**. When exposed through an MCP server and loaded into Claude Code and the OpenAI Codex desktop application, the system achieved Recall@100 of 0.958 and 0.967, respectively, on representative development-set concepts, with corresponding AP@100 values of 0.874 and 0.898. These case studies indicate that the tool interface and skill are portable to end-user agentic environments.
▪ **Independent test set**. On the silver benchmark of 457 concept-vocabulary pairs from 15 AD/ADRD target trial emulation studies, the best configuration achieved overall Recall@100 of 0.835 and AP@100 of 0.786. Recall@100 by vocabulary was 0.934 for ATC, 0.778 for ICD-10-CM, 0.738 for ICD-9-CM, 0.765 for ICD-10-PCS, and 0.832 for ICD-9-Proc. The drop relative to the curated development set is expected: published codelists embed study-specific scope decisions and local conventions that are not fully recoverable from a short concept description alone and the Jaccard index across papers reporting the same concept is only 0.760.

**Table 4.** Generalizability evaluation with OMOP Vocabulary Tools + Extension Layer and Progressive Guidance.

| Settings | Dataset | Agentic Application | Model | Recall@100 | AP@100 |
| --- | --- | --- | --- | --- | --- |
| Cross-model | Development Set | LangChain | GPT-5-mini | 0.965 | 0.875 |
|  |  |  | Qwen 3.5 397B A17B | 0.921 | 0.851 |
| Cross-application |  | Claude Code | Claude Sonnet 4.6 | 0.958 | 0.874 |
|  |  | Codex | GPT-5.3-Codex | 0.967 | 0.898 |
| Independent Testing | Independent test set | LangChain | GPT-5-mini | 0.835 | 0.786 |

## Discussion

This study demonstrates that OMOP concept-set authoring is best framed as a knowledge-guided agentic task rather than as a pure retrieval or ranking problem. The largest performance gains came from structured tools that transform raw vocabulary access into operations aligned with authoring workflows: search, expansion, mapping, validation, and refinement. Although Vanilla SQL exposed the same underlying data, it forced the agent to formulate every query and traversal step itself, leading to avoidable errors. PHOEBE 2.0 performed less well in our setup not because its underlying signals are necessarily uninformative, but because its interface was designed for interactive human review rather than autonomous multi-step execution.

A second major finding is that procedural knowledge matters, and its delivery strategy matters nearly as much as its content. Under the best tool configuration, Minimal Guidance already performed strongly, highlighting the importance of well-designed tools. The remaining gains came from helping the agent choose and sequence those tools for more complex concepts. Progressive Guidance outperformed Static Full Guidance while using fewer tokens and less time, suggesting that selective delivery of procedural knowledge reduces prompt dilution and keeps relevant guidance salient. In practical terms, the contribution did not come from the mere presence of a skill artifact; it came from selectively delivering procedural knowledge about concept decomposition, domain-specific retrieval routes, exclusion logic, validation, and stopping criteria. More generally, the findings support a platform-agnostic design in which an agentic controller is paired with structured tools and targeted procedural knowledge. In concept-set authoring, this combination is necessary because the task requires planning, vocabulary-specific tool selection, expert judgment about exclusions, and iterative refinement.

Concept-level and cohort-level evaluation revealed an important measurement nuance. Cohort metrics were consistently higher than concept-level recall because patient identification is dominated by a small number of high-prevalence codes. Concept-level retrieval therefore remains necessary for judging vocabulary coverage, but it may overstate the practical effect of missing rare concepts on downstream patient selection. Reporting both levels of evaluation gives a more realistic view of authoring quality and should become standard practice for future evaluations of automated concept-set authoring systems.

The independent silver benchmark highlights a second ceiling imposed by the task itself. Published codelists for the same named concept often differ because phenotype boundaries are study dependent^3,25^. When the input is only a short natural-language label, some inclusion and exclusion decisions are underdetermined. The system may therefore disagree with a specific reference set without being clinically unreasonable. Future systems should combine automated retrieval with either structured input templates or interactive clarification for boundary-ambiguous concepts, and should further formalize validation rules for vocabulary membership, domain consistency, and exclusion logic.

This study has several limitations. First, evaluation was centered on AD/ADRD, so broader generalization to domains with different vocabulary structures remains to be demonstrated. Second, the PHOEBE comparison reflects autonomous use under our generic agent design rather than a PHOEBE-optimized agent strategy. Our results therefore represent performance under one agentic operating mode, not PHOEBE’s theoretical upper bound. Third, the silver benchmark inherits between-study variability in published codelists, which limits its ability to distinguish true system errors from legitimate design differences. Fourth, we did not conduct end-user studies to evaluate whether the system reduces expert review time, affects trust, or improves collaborative authoring workflows in practice.

Several directions for future work follow naturally. Future work should extend evaluation across additional disease domains, design agent strategies tailored to PHOEBE’s recommender interface, add deterministic validators for critical checks, and test human-in-the-loop workflows in which the agent proposes a candidate concept set and explicitly flags boundary-ambiguous descendants for adjudication. These extensions would help clarify when autonomous authoring is sufficient and when expert clarification remains essential.

## Conclusion

We presented a knowledge-guided agentic framework for OMOP concept-set authoring from natural language descriptions. Our findings show that structured, task-specific tools contribute more to reliable authoring than unrestricted raw data access, and that targeted external resources help close known coverage gaps. Procedural knowledge provides additional value, particularly for concepts that require exclusions, multi-hop retrieval, or cross-resource reasoning, but that value is maximized when the knowledge is delivered selectively rather than all at once. This procedural knowledge can be packaged as a modular skill artifact, and more broadly, our findings support an architectural pattern that combines agent controller with structured tools and targeted procedural knowledge. Together, these components enable accurate, scalable, and reproducible OMOP concept-set authoring for clinical informatics applications.

## Data Availability

All data produced in the present study are available upon reasonable request to the authors.

## Acknowledgements

This work was supported in part by NIH grants R01AG084236. The content is solely the responsibility of the authors and does not necessarily represent the official views of the NIH.

